# Hyperbolic weight-height correlation in children and adolescents: models and implications for obesity diagnosis

**DOI:** 10.1101/2025.08.16.25333345

**Authors:** Taukim Xu, Xu Li, Wenbin Liu, Na Zhao, Qian Wu

**Author notes:** Correspondence author (Taukim Xu;).

## Abstract

Many weight-height (Wt-Ht) models were proposed since the derivation of the “Quetelet Index”, though their credibility remains elusive. This raises an interesting question about the presence of numerous Wt-Ht models and reveals that our knowledge about the Wt-Ht correlation is still far from its essence. We identified a strong linear correlation between Wt and Wt*Ht^c^ (c=-2∼2), based on data from 359,049 participants aged 1 to 21 years, recruited from China, Japan, South Korea, Slovakia, the USA, and Bangladesh. Then, we established a statistically-robust Wt-Ht model, which is expressed as Wt=a+b*Wt*Ht^c^ (Eq. 1) or 1=a/Wt+b*Ht^c^ (Eq. 2), through linear regression. When c=1 or -1, Eq. 2 is a standard hyperbolic function, which proves that Wt is hyperbolically correlated with Ht. The coefficients a and b are sex-, age-, and geography-specific constants. As the exponent c approaches 0, the correlation between Wt and Wt*Ht^c^, along with the standardized Wt-Ht index (sWHI) (a/Wt+b*Ht^c^ in Eq. 2), approaches 1. Further, we incorporated total fat to determine the implications of our model in obesity diagnosis. When c=1, the sWHI (a/Wt+b*Ht) is capable of screening abnormal body fat percentage (BFP) based on deviations from the Wt-Ht equilibrium (sWHI=1). BMI demonstrates advantages over other anthropometric indexes in screening abnormal BFP; nonetheless, their performances are largely similar. This is probably attributed to the strong linear correlation between Wt and Wt*Ht^c^. Overall, the hyperbolic Wt-Ht model reveals the nature of Wt-Ht correlation and offers key insights into obesity diagnosis using anthropometric indexes.

## Main

Weight (Wt) and height (Ht) are two fundamental physical traits, reflecting the health condition and social status [C1,C2,C3,C4,C5,C6,C7,C8]; however, they are heterogeneous across populations [C4,C9,C10,C11,C12]. Numerous factors, such as environmental influences, genetics, and nutritional status [C4,C10,C11,C13,C14,C15,C16,C17], affect these easily measurable anthropometric variables. Therefore, establishing a reliable mathematical model to explain the relationship between Wt and Ht is a critical and foundational area of human research [C1,C2,C6,C18,C19].

Many Wt-Ht models have been proposed and roughly categorized into linear or nonlinear models [C1,C2,C6,C20,C21,C22,C23]. The linear model assumes that Wt increases as a linear function of Ht. Advocates of the linear model include the Wt/Ht tables, published by the Metropolitan Life Insurance Company based on extensive life insurance data [C6,C24], and the subsequent ideal body weight (IBW) concept [C20], calculated via the Wt/Ht tables [C21]. The IBW is used to predict the ideal or desirable Wt for individuals with the lowest mortality risk. On the other hand, the Benn-type indices (Wt/Ht^β^) were proposed [C2,C25]—when β=1, it is the linear weight-height model—to capture the nonlinear correlation between Wt and Ht and to make the index uncorrelated with Ht. The body mass index (BMI) is the most widely accepted Benn-type index, when β=2. It is used in obesity diagnosis [C1,C5,C7,C18], despite its limitations in distinguishing fat and lean body mass [C1,C5,C6].

Overall, many Wt-Ht models (Wt^1/3^/Ht, Wt/Ht, Wt/Ht^2^, Wt/Ht^3^, etc.) have been developed, each appearing reasonable and valid [C18,C26,C27]. This raises an interesting question about the presence of numerous Wt-Ht models. In this study, we aimed to investigate the nature of the Wt-Ht correlation and determine the implications of our findings for obesity diagnosis.

## Results

### Datasets

Two data sources were used for analysis. The first set—collected by our team— facilitated the initial discovery of a strong linear correlation between Wt and Wt*Ht^c^ (c=-2∼2). The second set—obtained from seven open-source datasets—was used to validate our findings and suggest implications for obesity diagnosis (Extended Data Fig. S1). Our dataset included 6,831 records, of which 6,226—including 2,823 females (45.3%) and aged 6 to 21 years—were used in the final analysis. The seven open-source datasets comprised data from 352,823 participants, including 157,824 females (44.7%) and aged 1 to 21 years, recruited from China, Japan, South Korea, Slovakia, the USA, and Bangladesh. Thus, the final analysis comprised 359,049 records, including data from 160,647 females (44.74%) and aged 1 to 21 years. These records were divided into two groups as follows: (1) comprising measured age, sex, height, and weight; (2) comprising measured total fat (measured by Dual-Energy X-ray Absorptiometry), age, sex, height, and weight (Extended Data Fig. S2).

### Strong linear correlation between Wt and Wt*Ht^c^

We observed an extremely strong linear correlation (r>0.97) between Wt and Wt*Ht (Fig. 1a) during a routine regression analysis, where Wt and Wt*Ht were included as independent variables to determine the maximum hand grip strength (mHGS) [C28]. A strong collinearity was observed between Wt and Wt*Ht. In comparison, no such strong linear correlation was observed between Ht and Ht*Wt (Fig. 1a). Further investigation suggested a similar strong linear correlation between Wt and Wt/Ht.

**Fig. 1:**
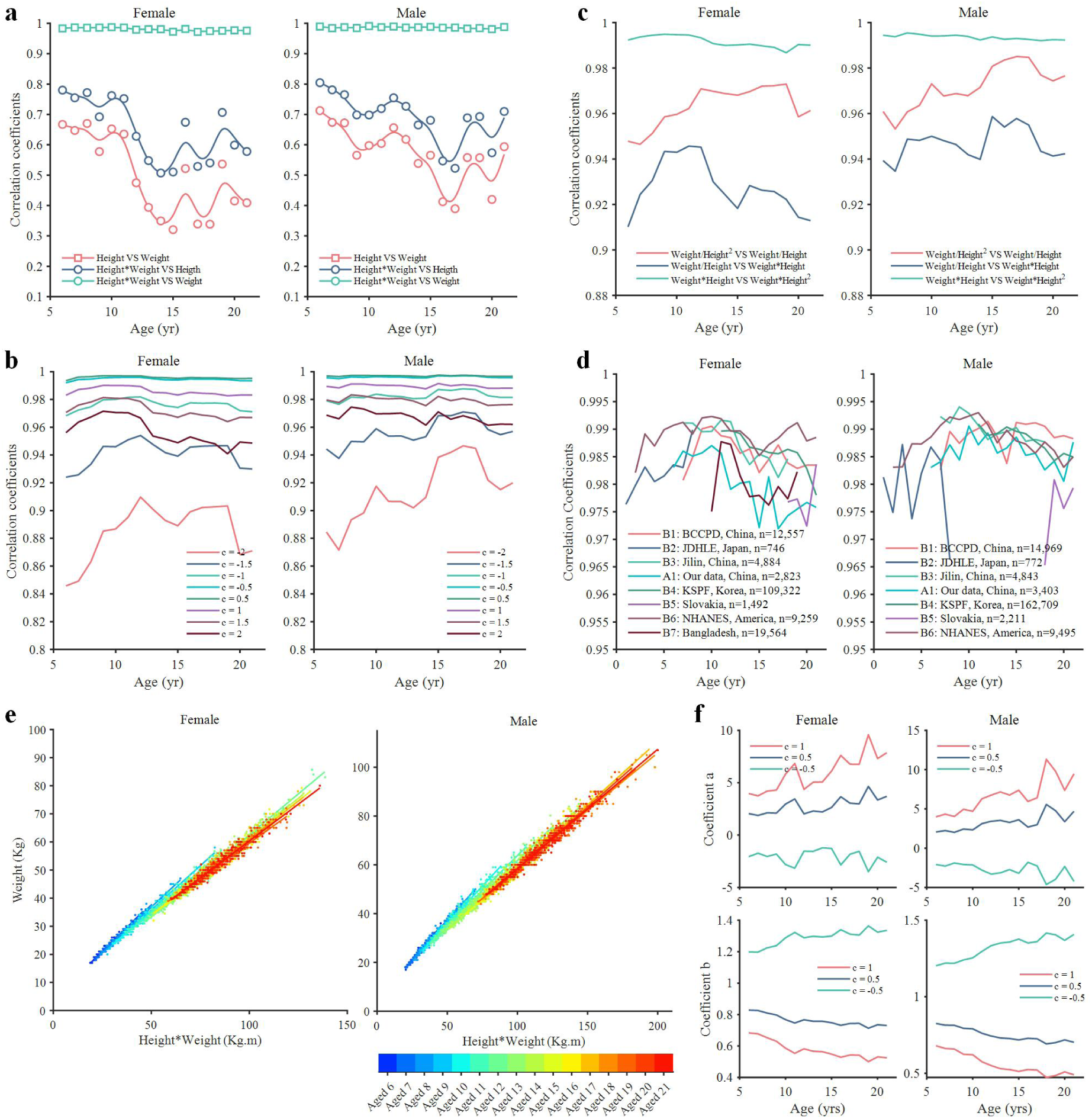
Strong linear correlation between Wt and Wt*Ht^c^ and the hyperbolic Wt-Ht model. **a,** Linear correlation coefficients between Wt and Ht, Wt*Ht and Ht, and Wt*Ht and Wt for A1: Our data. **b,** Linear correlation coefficients between Wt and Wt*Ht^c^ when exponent c ranges from -2 to 2 for all Chinese datasets. **c,** Linear correlation coefficients between Wt/Ht^2^ and Wt/Ht, Wt/Ht and Wt*Ht, and Wt*Ht and Wt*Ht^2^ for all Chinese datasets. **d,** Linear correlation coefficients between Wt and Wt*Ht for all datasets. **e,** Linear regression of the hyperbolic Wt-Ht model when exponent c=1 for A1: Our data. **f,** Variations of coefficients a and b with age when the exponent c=1, 0.5, and -0.5 for A1: Our data. Wt = weight; Ht = height; BCCPDS = Body Composition of Chinese People Data Set; JDHLE = Japanese Database for Human Life Engineering; KSPF = Korea Sports Promotion Foundation; NHANES = National Health and Nutrition Examination Survey

Thus, we constructed a more generalized term—Wt*Ht^c^—to examine the linear correlation between Wt and Wt*Ht^c^ as the exponent c varied from -2 to 2. A strong linear correlation (r>0.92) was observed, except for c=-2 (r>0.82) (Fig. 1b). When c approached 0, the correlation coefficient approached 1. Simultaneously, a strong linear correlation (r>0.91) was observed within the Wt*Ht^c^ indexes (c=-2 vs. c=-1, c=- 1 vs. c=1, and c=1 vs. c=2) (Fig. 1c). When c=-2, Wt*Ht^c^ represents the BMI model; in contrast, when c=-1, Wt*Ht^c^ represents the linear Wt-Ht model. Notably, the extremely strong linear correlation (r>0.97) between Wt and Wt*Ht was verified across all datasets (Fig. 1d).

### Hyperbolic Wt-Ht model

Based on the strong linear correlation between Wt and Wt*Ht^c^, we established a linear regression model (Fig. 1e and Extended Data Fig. S3) [C29]. In this model, Wt served as the dependent variable, whereas Wt*Ht^c^ served as the independent variable. To simplify the formula, the error random variable was excluded as shown below:

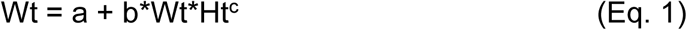

and its transformation:

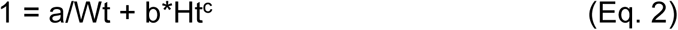

Here, the coefficients a and b are sex-, age-, and geography-specific constants, obtained through linear regression of Eq. 1. Wt should be greater than 0 and the recommended range for c is -2 to 2 to achieve a good regression effect. When c=1 or -1, Eq. 2 is a standard hyperbolic function, which proves that Wt is hyperbolically correlated with Ht.

Eq. 2 suggests that human Wt and Ht can be normalized to 1 using specific coefficients a and b. This resulted in the definition of the standardized Wt-Ht index (sWHI), expressed as a/Wt+b*Ht^c^. sWHI shows a bell-shaped distribution and exhibits good normality across all datasets (Fig. 2a,b and Extended Data Figs. S3, S4, and S5). Additionally, when c approaches 0, the range between the 5^th^ and 95^th^ percentiles of sWHI narrows linearly (Fig. 2c,d and Extended Data Fig. S4). Eqs. 1 and 2 can be transformed to predict Ht or Wt. When c=1, the difference between the real and predicted Wt and Ht (represented by Wr-Wp and Hr-Hp) shows bell-shaped distributions across all Chinese and American datasets (Fig. 2e,f). Interestingly, compared with the Hr-Hp curve, the Wr-Wp curve is sharper and shows a wider distribution. Overall, the strong linear correlation between Wt and Wt*Ht and the bell- shaped curves confirm the statistical validity of the hyperbolic correlation between Wt and Ht.

**Fig. 2:**
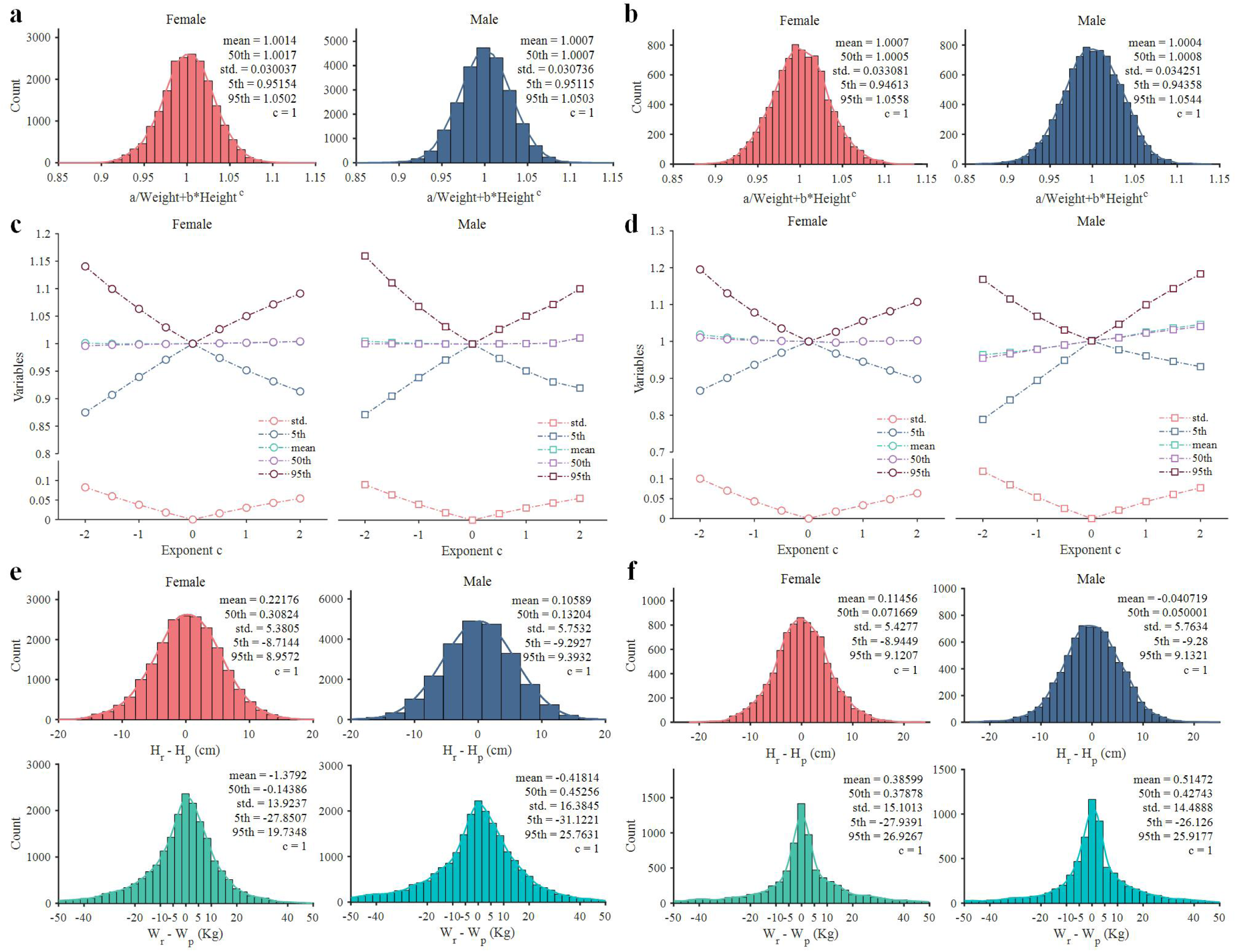
Bell-shaped distribution curve of sWHI, Hr-Hp, and Wr-Wp when the exponent c=1. **a,b**, Distribution diagrams of sWHI for all Chinese (including A1, B1, and B3) (**a**), and American (including B6) (**b**) datasets based on the quadratic polynomial regression models of coefficients a and b. **c,d**, Standard deviation, mean, and 5^th^, 50^th^, and 95^th^ percentiles of sWHI change linearly when the exponent c ranges from -2 to 2 for all Chinese (**c**) and American (**d**) datasets. **e,f**, Distribution diagrams of Hr-Hp and Wr-Wp for all Chinese (**e**) and American (**f**) datasets. Mean, standard deviation, and 5^th^, 50^th^, and 95^th^ percentiles are shown for all sWHI, Hr-Hp, and Wr-Wp distributions. sWHI = standardized weight-height index; Hr = real height; Hp = predicted height; Wr = real weight; and Wp = predicted weight; Wt = weight; Ht = height; std.= standard deviation

### Sex-, age-, and geography-specific coefficients

Linear regression analysis indicated the changing patterns of coefficients a and b in Eqs. 1 and 2 concerning age, sex, geography, and the exponent c. Exponent c affects these coefficients, which continuously vary with age (Fig. 1f). When c=1, coefficient a increases with age, whereas coefficient b decreases with age (Fig. 3a), and both display nonlinear trajectories for females and males. Quadratic polynomial models were, therefore, established to describe these changes for all (Fig. 3b), Chinese (Fig. 3c), and American (Fig. 3d) datasets. Quadratic polynomial regression exhibited lower determination coefficient for coefficient a (R^2^= 0.24536–0.48632) than that for coefficient b (R^2^= 0.82602–0.96209). Differences in R^2^ may explain the sharper and wider Wr-Wp distribution at both sides, compared with Hr-Hp (Fig. 2e, f). Extended Data Fig. S6 shows how coefficients a and b vary with age when c=-1, -0.5, and 0.5 for all Chinese (Extended Data Fig. S6a,c,e) and American (Extended Data Fig. S6b,d,f) datasets, respectively.

**Fig. 3:**
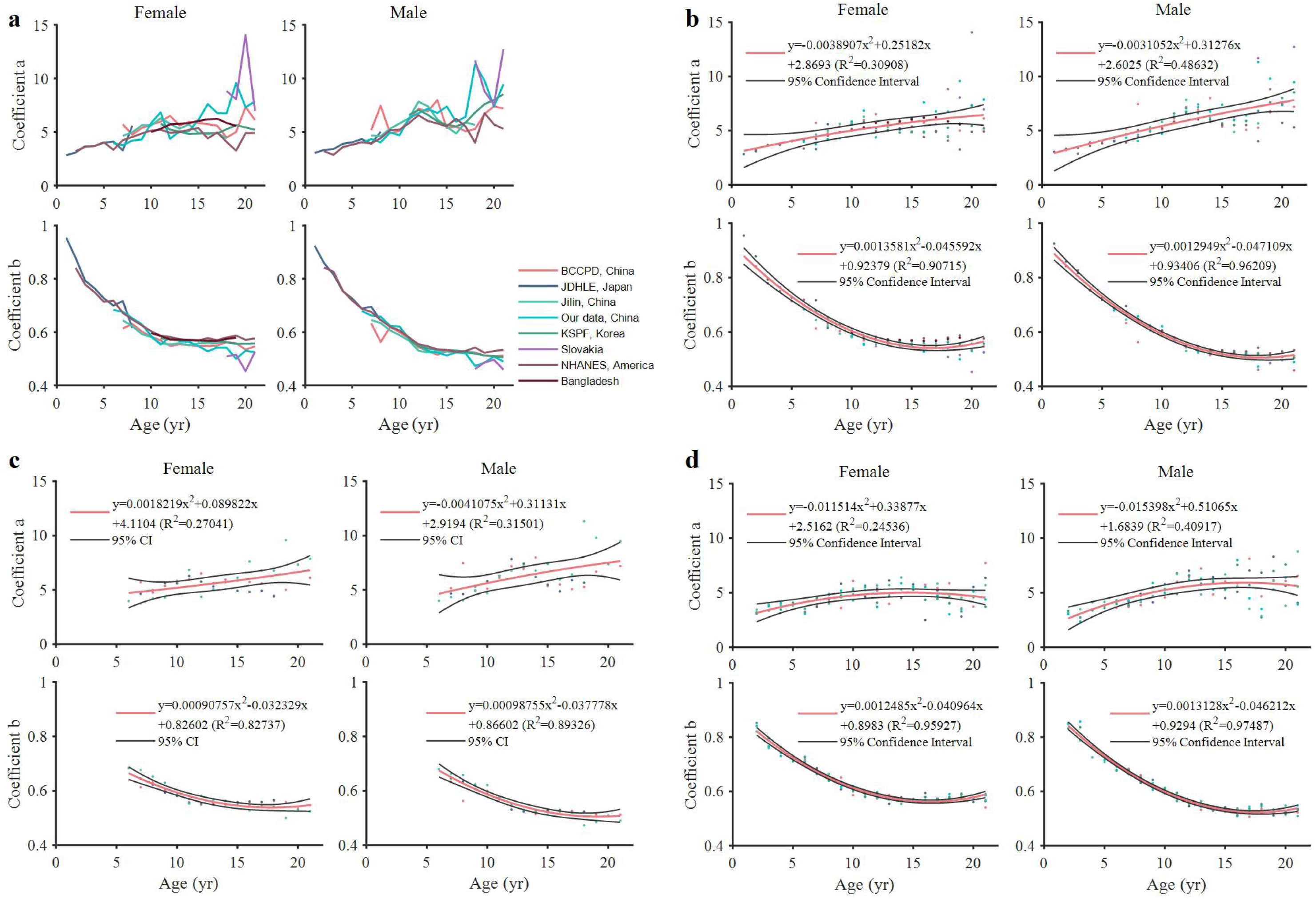
Sex-, age-, and geography-specific coefficients a and b when the exponent c=1. **a,** Variation of coefficients a and b with age for all datasets. **b-d,** Quadratic polynomial regression models of coefficients a and b for all (**b**), Chinese (including A1, B1, and B3) (**c**), and American (including B6) (**d**) datasets. For **a** and **b,** all NHANES datasets were integrated for analysis, whereas for **d,** data from NHANES 2011-2012, NHANES 2013-2014, NHANES 2015-2016, NHANES 2017-2018, and NHANES 2017-March 2020 were analysed separately. BCCPDS = Body Composition of Chinese People Data Set; JDHLE = Japanese Database for Human Life Engineering; KSPF = Korea Sports Promotion Foundation; NHANES = National Health and Nutrition Examination Survey; CI = confidence interval; R^2^ = determination coefficient in regression analysis

### sWHI performance in obesity diagnosis

The sWHI is normally distributed around 1 (Fig. 2a,b and Extended Data Figs. S3, S4, and S5). Therefore, an interesting question is that what does it mean if the sWHI deviates too much from 1. According to Eq. 2, when c=1, participants who are light and tall, suggesting a high risk of being underweight, demonstrate significantly higher sWHI values. Therefore, we explored the applicability of sWHI in obesity diagnosis when c=1 (Fig. 4). First, we calculated the sWHI for each age group and merged them using quadratic polynomial models of coefficients a and b. Second, based on the sWHI percentiles, data were categorized into three groups (sWHI≤5^th^, 5^th^<sWHI<95^th^, and sWHI≥95^th^) and BMI distribution of each group is displayed in box plots. The BMI and body fat percentage (BFP), calculated as total fat/Wt, were considered because BMI is the most commonly used criteria for obesity diagnosis [C18] and the World Health Organization (WHO) defines obesity as abnormal or excessive fat accumulation, which can be quantized using BFP, that impairs health [C7].

**Fig. 4:**
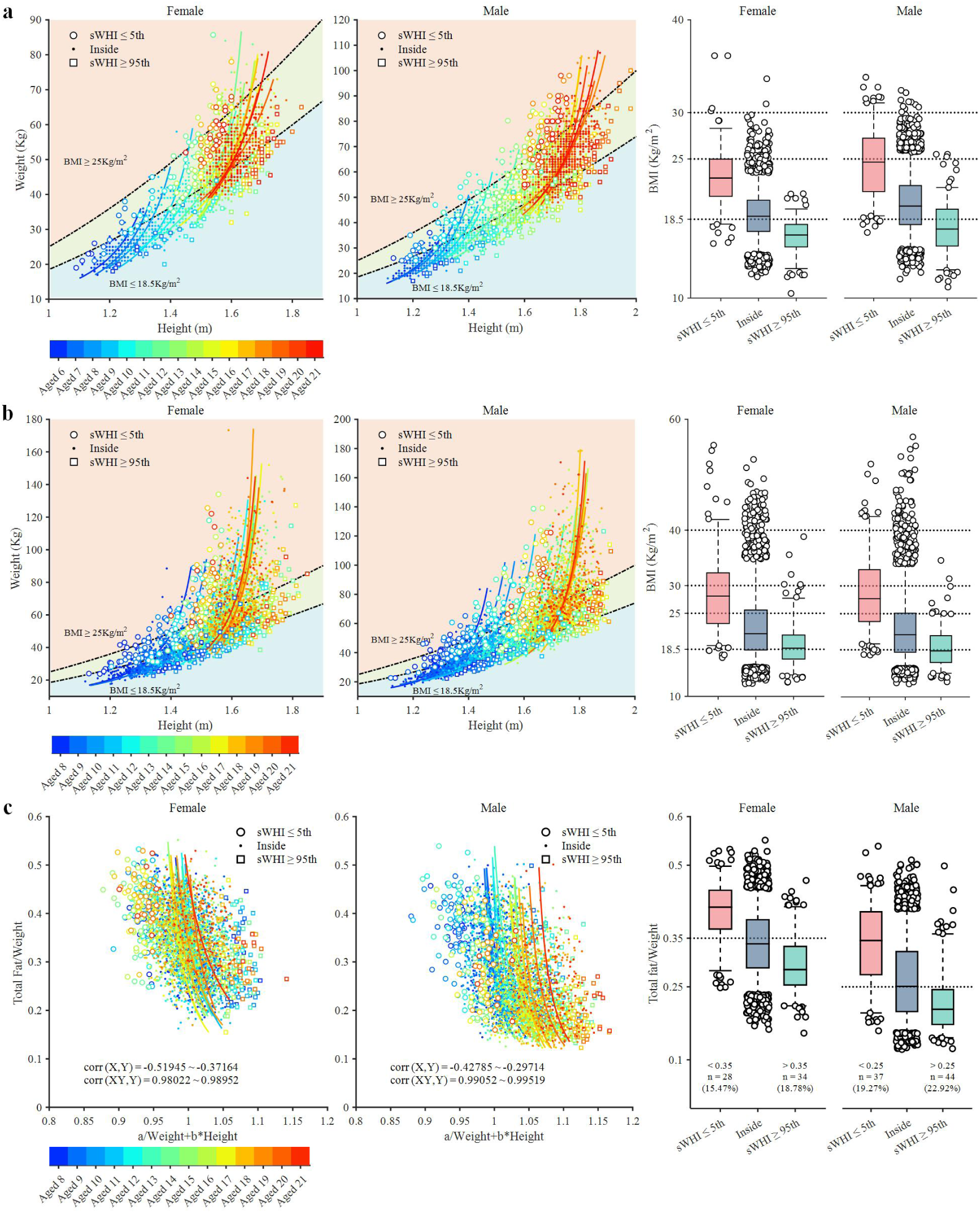
sWHI performance in screening abnormal BFP when the exponent c=1. **a-b,** Comparison between the hyperbolic (c=1) and BMI models for Chinese (**a**) and American (**b**) datasets. The first two panels show scatter plots of Wt and Ht with data stratified by sex, age, and percentiles (5^th^ and 95^th^) of sWHI. The solid line represents the hyperbolic Wt-Ht model (Eq. 2). The frame is divided into three areas as follows: BMI≥25 Kg/m^2^, 18.5 Kg/m^2^<BMI<25 Kg/m^2^, and BMI≤18.5 Kg/m^2^. Data for sWHI≤5^th^ are marked with circles, whereas those for sWHI≥95^th^ are marked with squares. The last panel shows box plots for BMI with data stratified by sex and sWHI percentiles. In each box, data outside the 5^th^ and 95^th^ BMI percentiles are plotted as black circles. **c,** sWHI performance in screening abnormal BFP. The first two panels show scatter plots of sWHI and BFP (Total fat/Weight) with data stratified by sex, age, and sWHI percentiles. Data for sWHI≤5^th^ are marked with circles, whereas those for sWHI≥95^th^ are marked with squares. sWHI*BFP shows an extremely high linear correlation with BFP. sWHI shows a low or moderate correlation with BFP. The last panel shows box plots for BFP with data stratified by sex and sWHI percentile. In each box, data outside the 5^th^ and 95^th^ BFP percentiles are plotted as black circles. BMI = Body Mass Index; sWHI = standardized weight-height index; BFP = body fat percentage; Wt = weight; Ht = height

For BMI (Fig. 4a,b), the BMI model with a fixed standard tended to classify younger participants as underweight (BMI≤18.5 kg/m^2^), with approximately all 6-year- olds falling into this category (Fig. 4a). Thus, the using of BMI for obesity diagnosis are age-dependent, and we grouped the data by age and stratified them by sWHI percentiles for each age group. After stratifying by sWHI, the median BMI in each sWHI group decreased with increasing sWHI. However, some participants in the sWHI≤5^th^ groups had BMI≤18 kg/m^2^, suggesting underweight rather than obesity.

Therefore, sWHI does not appear to be a reliable obesity indicator when using BMI as the standard. Furthermore, we incorporated total fat into the comparison and analyzed BFP distribution across sWHI groups (Fig. 4c). We observed a strong linear correlation between BFP and BFP*sWHI, confirming the hyperbolic correlation between sWHI and BFP. In the sWHI≤5^th^ group, most females (over 85%) had BFP≥0.35 and most males (over 77%) had BFP≥0.25. Thus, the sWHI can be used to screen high or low BFP based on deviations from the Wt-Ht equilibrium (sWHI=1). Nonetheless, some participants had BFP>0.35 or 0.25 in the sWHI≥95^th^ and 5^th^<sWHI<95^th^ groups. Therefore, excessive deviations from the Wt-Ht equilibrium are not the sole contributors to abnormal BFP.

### Wt*Ht^c^ index performance in screening low or high BFP

We stratified the data using Wt*Ht^c^ index percentiles, including the Wt, Wt/Ht, BMI, and Wt*Ht when the exponent c=0, -1, -2, and 1, respectively, and examined BFP distribution within each group (Fig. 5). Notably, a strong linear correlation (r = 0.82288–0.96051) was observed between Wt*Ht^c^ and BFP*Wt*Ht^c^ (Fig. 5a).

**Fig. 5:**
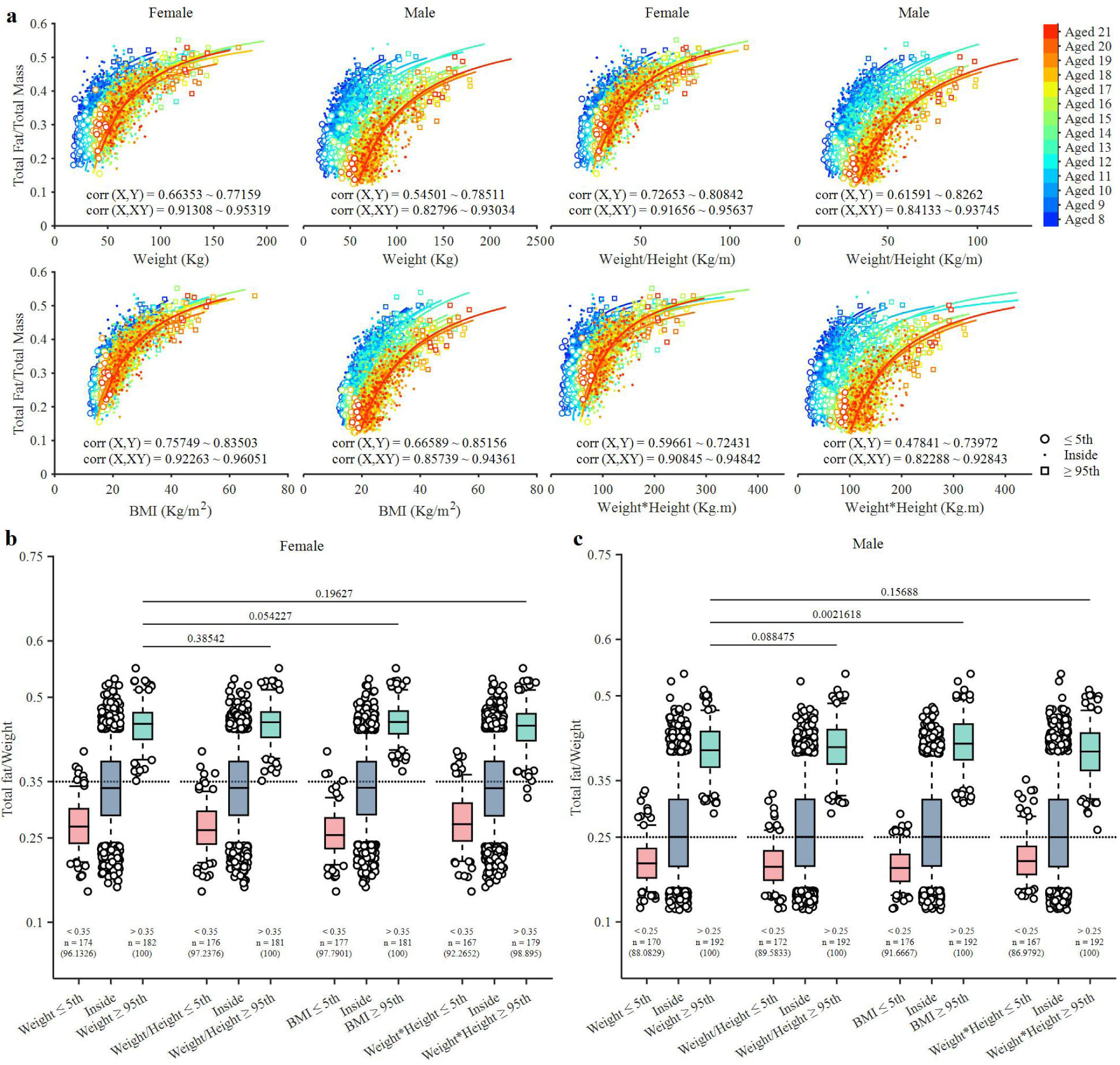
Wt*Ht^c^ index performance in screening abnormal BFP when the exponent c=0,-1,-2, and 1. **a,** Scatter plots between BFP and Wt*Ht^c^ with data stratified by sex, age, and 5^th^ and 95^th^ percentiles of Wt*Ht^c^. Data of Wt*Ht^c^≤5^th^ are marked with circles, whereas those of Wt*Ht^c^≥95^th^ are marked with squares. The solid line represents the hyperbolic model, since there is a strong linear correlation between X and X*Y. **b,c**, Box plots for BFP with data stratified by sex and 5^th^ and 95^th^ percentiles of Wt*Ht^c^ for females (**b**) and males (**c**). In each box, data outside the 5^th^ and 95^th^ BFP percentiles are plotted as black circles. For each category, the number and proportion of BFP<0.35 or >0.35 for females and BFP<0.25 or >0.25 for males are shown. BMI = Body Mass Index; BFP = Body Fat Percentage; Wt = weight; Ht = height

Furthermore, when c=0, -1, -2, and 1, groups with Wt*Ht^c^≥95^th^ demonstrated excellent performance in screening high BFP, with the BFP proportion >0.35 and >0.25 being 98.895% in females and 100% in males, respectively (Fig. 5b,c). BMI displayed a minimal advantage over the other three variables in screening high BFP. Visually, medians and percentiles between the groups were almost identical, and among females, no difference (p=0.054227 in student’s t-test or Mann–Whitney U test) was observed between Wt and BMI for the Wt*Ht^c^≥95^th^ group (Fig. 5b). This finding may be attributed to the strong linear correlation between Wt and Wt*Ht^c^ (Fig. 1b) and the capacity of Wt*Ht^c^ indexes to screen for abnormal BFP based on their strong correlation with Wt. Similarly, in the Wt*Ht^c^≤5^th^ and 5^th^<Wt*Ht^c^<95^th^ groups, some participants had BFP>0.35 or 0.25. Therefore, the causes of high BFP are complex and diverse [C18], including deviations in anthropometric variables.

## Discussion

Based on a large sample of data from multiple countries and regions, we identified a hyperbolic correlation between Wt and Ht in children and adolescents. The mathematical relationships between Wt and Ht take various hyperbolic forms as the exponent c in Eq. 2 changes. This finding well explains why there are so many Wt-Ht models. Actually, these Wt-Ht models are statistically equivalent with mathematical nuances, and both linear and nonlinear models [C2,C6] approximate the real Wt-Ht correlation. This equivalence is emphasized by the facts that all the Wt*Ht^c^ indexes are linearly correlated to with Wt, and that there is a good strong linear correlation between Wt/Ht and BMI (Fig. 1a-d).

The hyperbolic correlation between Wt and Ht can be explained mathematically. In Eq. 1, when c approaches 0, Ht^c^ approaches to 1. Thus, Wt*Ht^c^ approaches Wt. This is why, when c approaches 0, the closer the correlation coefficient between Wt and Wt*Ht^c^ approaches 1 and the gap between the 5^th^ and 95^th^ sWHI percentiles narrows linearly (Fig. 1b and Fig. 2c,d). Actually, when the exponent c ranges from -0.1 to 0.1, a strong linear correlation (r>0.778152) is observed between Ht and Ht*Wt^c^ (Extended Data Fig. S8). However, the hyperbolic Wt-Ht model is expressed as Wt≈Wt*Ht^c^ rather than Ht≈Ht*Wt^c^ because this formulation (Eq. 2) encompasses a wider range of c and a good normal distribution of Hr-Hp and Wr-Wp (Fig. 2a,b vs. Extended Data Fig. S8e,f). This mathematical strategy has wider applicability in establishing complex correlations between variables. For example, we observed a similar hyperbolic correlation between mHGS and Ht (mHGS≈mHGS*Ht), though it is beyond the scope of this article. Therefore, such correlations are probably widespread across biological traits and even in nature.

The hyperbolic Wt-Ht correlation is statistically robust and universal for these reasons:

1. The extremely strong linear correlation (r>0.97) between Wt and Wt*Ht has been validated on a large, diverse dataset.
2. The distribution of sWHI, Hr-Hp, and Wr-Wp formed bell-shaped curves, suggesting Gaussian or normal distributions.

sWHI is symmetrically distributed around 1, even upon using the quadratic polynomial regression models with poor determination coefficient (R^2^) (Fig. 3b,c,d). In contrast, BMI displays an asymmetrical distribution and skews right toward a higher Wt to Ht^2^ ratio [C6,C30]. The normal distribution of sWHI indicates an equilibrium between Wt and Ht, where sWHI=1. Deviations from this equilibrium are correlated with abnormal BFP. Furthermore, quadratic polynomial regression models for coefficients a and b, along with Eqs. 1 and 2, provide key tools for clinical staff to estimate the Wt in a child or adolescent based on Ht in critical care scenarios [C31,C32].

The BMI model is widely used as a preliminary screening tool for obesity [C7,C18,C33]. Supporters of BMI-based obesity diagnosis argue that BMI is strongly correlated with body fat [C5,C34], consistent with the WHO’s definition of obesity as the abnormal and/or excessive accumulation of body fat impairing health [C7,C18]. However, weight is strongly correlated with body fat (Extended Data Fig. S9a). Considering it does not require additional calculations, it is reasonable to consider using Wt for obesity diagnosis directly. A compelling argument supporting the use of BMI is that BMI is independent of Ht [C2]. However, BMI does not show a substantial advantage over Wt in identifying individuals with high BFP based on our findings (Fig. 5b,c). A plausible explanation for the feasibility of anthropometric indexes for obesity diagnosis is their strong correlation with Wt. This is because Wt comprises body fat and increased body fat leads to increased body Wt. Additionally, these anthropometric indexes cannot be used as the sole criteria for obesity diagnosis [C18]. All groups with 5^th^<sWHI<95^th^, sWHI≥95^th^, Wt*Ht^c^≤5^th^, and 5^th^<Wt*Ht^c^<95^th^ comprised participants with BFP>0.35 or 0.25 (Fig. 4c and Fig. 5b,c). Thus, Wt-Ht imbalance does not seem to be the only contributor of abnormal BFP and a substantial Wt-Ht imbalance increases the risk of abnormal BFP. That is the reasons behind abnormal BFP are complex, with anthropometry being one of the factors. At the same time, our analysis emphasizes age-stratified anthropometric indexes and determining obesity diagnostic criteria based on percentiles within each age group. The BMI model with a fixed standard tends to classify younger participants as underweight (Fig. 4a, b), indicating its unfeasibility for diagnosing childhood obesity [C19,C26].

Several limitations of this study should be acknowledged. We collected global data to confirm the universality of the hyperbolic Wt-Ht correlation; nonetheless, data from Africa and Oceania were lacking. Therefore, global variations in coefficients a and b in Eq. 1 or Eq. 2 remained elusive. Additional data-based evidence is required to establish the global applicability of the hyperbolic Wt-Ht model. However, the quadratic polynomial models for coefficients a and b (Fig. 3c, d) were based on large- sample nationwide data, from China and the USA. Thus, combined with Eq. 1 or Eq. 2, these models provide key insights into Wt and Ht management for these populations. Meanwhile, the coefficient a does not follow a quadratic form with age, necessitating alternative forms of fitting model to improve the goodness of fit.

However, we used the quadratic polynomial model for the purpose of improving data comparability across different countries and regions. Besides, this study primarily focused on children and adolescents, with limited data on adults [C35]. Likewise, additional data from older adults (aged >30 years) are required to confirm our findings across all age groups. Moreover, future research should explore the correlation between the sWHI index and other health risks, such as mortality [C36,C37,C38,C39]. This is because Wt-Ht imbalance probably results in adverse health outcomes. However, despite these limitations, our findings are novel, substantial, and groundbreaking.

Over all, we identified a statistically robust hyperbolic correlation between Wt and Ht in children and adolescents, counteracting existing knowledge about Wt-Ht correlations. After the derivation of the “Quetelet Index,” the hyperbolic Wt-Ht model provides a novel theoretical tool to uncover the nature of Wt-Ht correlation and explain the feasibility of these anthropometric indexes for obesity diagnosis. Wt-Ht indexes (Wt*Ht^c^) screen for abnormal BFP probably based on their strong correlation with body Wt. Besides, all anthropometric indexes perform similarly in screening for abnormal BFP and anthropometry is one of the factors that lead to abnormal BFP. Therefore, we need to be more rational about using Wt-Ht indexes to screen for obesity.

## Methods

### Study design

We started by establishing an empirical model for predicting maximum hand grip strength (mHGS), a crucial health indicator [C40], based on demographic and physical parameters [C28]. To this end, we initiated a long-term program in April 2022 to collect large sample data, including age, sex, handedness, height (Ht), weight (Wt), forearm circumference, and mHGS, from healthy Chinese individuals. Presently, this program focuses on only school students; therefore, participants aged 6 to 21 years were included in the analysis. We did not distinguish between rural and urban participants because the advantages of urban living for growth and development are diminishing in both children and adolescents [C11]. The exclusion criteria included pregnancy, pain or restriction of movement in a hand or arm, neuromuscular disease, generalized bone disease, aneuploidy, any condition severely interfering with normal growth or requiring hormonal supplementation, and unwillingness to participate. Additionally, we searched for open-source datasets to confirm our findings, including data from participants aged 1 to 21 years.

### Data source and ethic matters

Extended Data Fig. S1 summarizes the data collection process, data inclusion and exclusion, and data flow. Two data sources were used for our analysis: (1) data collected by our team and (2) data obtained from seven open-source datasets. The “Data and code availability” section summarizes raw data.

For the data collected by our team, all participants were school students from six institutes, including one primary school (Yucai Primary School in Tongnan District of Chongqing), one junior high school (Sankou Junior High School in Jiangjin District of Chongqing), one high school (Tongnan No.1 Middle School in Tongnan District of Chongqing), and three universities (Chongqing University of Posts and Telecommunications, Chongqing College of International Business and Economics, and Chongqing University of Arts and Science), in Chongqing, southwest China. All participants provided written informed consent. For participants aged under 18 years, verbal consent was obtained from their parents or guardians. Physical education teachers and researchers ensured participant safety during all measurements. This study was approved by the Institutional Review Board of the People’s Hospital of Chongqing Banan District (approval number BNLL-KY-2023-004).

The second data source comprised seven open-source datasets as given below:

1. B1: Body Composition of Chinese People Data Set (BCCPDS) [C41], a nationwide study with data from Beijing, Guangdong, and other locations. Age, sex, Ht, and Wt of 27,526 participants (including 12,557 females) aged from 7 to 21 years were selected from this data-set.
2. B2: Japanese Database for Human Life Engineering (JDHLE) [C42], a regional study with data from Osaka Prefecture, Kyoto Prefecture, and other locations. Age, sex, Ht, and Wt of 1,518 participants (including 746 females) aged 1 to 8 years were selected from this data-set.
3. B3: Data from Wang, et al. published on Figshare [C43], including age, sex, Ht, and Wt of 9,713 participants (including 4,884 females) aged 7 to 18 years, recruited from Jilin, China.
4. B4: Korea Sports Promotion Foundation (KSPF) [C44], a nationwide study providing insights into sports, physical fitness data, health, and happiness in South Korea. Age, sex, Ht, and Wt of 272,031 participants (including 109,322 females) aged 11 to 21 years were selected from this data-set.
5. B5: Data from Miloš et al. [C45], including age, sex, Ht, and Wt of 3,703 participants (including 1,492 females) aged 18 to 21 years in Slovakia.
6. B6: National Health and Nutrition Examination Survey (NHANES) [C46], a USA-based program assessing the health and nutritional status of adults and children. It examines a nationally representative sample of approximately 5,000 persons each year. This study included the data of the NHANES 2011-2012, NHANES 2013-2014, NHANES 2015-2016, NHANES 2017-2018, and NHANES 2017-March 2020. Age, sex, Ht, and Wt of 18,754 participants (including 9,259 females) aged 2 to 21 years, and total fat, age, sex, Ht, and Wt of 7,477 participants (including 3,599 females) aged 8 to 21 years were selected from this data-set.
7. B7: Data from Akib et al. [C47], including age, sex, Ht, and Wt of 19,564 female aged 10 to 19 years in Bangladesh.

Because identifiable information was removed from raw data of the seven open- source datasets, we did not request additional ethical review for this part of data.

### Data inclusion and exclusion

Our team collected 6,831 records from school students aged from 6 to 25 years. We excluded participants aged over 21 years (N=605, 8.86%) because of insufficient sample sizes. The seven open-source datasets yielded 352,809 records from participants aged 1 to 21 years. All records were included in the analysis. Therefore, 359,035 records, including 160,647 females (44.74%), with measured age, sex, Ht, and Wt were analyzed to confirm the hyperbolic Wt-Ht correlations. Additionally, total fat (measured by Dual-Energy X-ray Absorptiometry) from the NHANES 2011-2012, NHANES 2013-2014, NHANES 2015-2016, and NHANES 2017-2018 datasets, obtained from 7,477 participants—including 3,599 females (48.1%)—were analyzed to investigate the implication of our findings for obesity diagnosis.

### Data processing

Extended Data Figure S2 illustrates the data flow. All data were labeled as two groups. The first group comprised 359,035 records with age, sex, Ht, and Wt measured, whereas the second group comprised 7,477 records with total fate, age, sex, Ht, and Wt measured. For the first group, data were stratified by age, sex, and country or region. Correlation analysis was conducted to identify the strong linear correlation between Wt and Wt*Ht^c^. Linear regression was utilized to develop the hyperbolic Wt-Ht model, yielding sex-, age-, and geography-specific coefficients a and b. Thereafter, bell-shaped distribution curve of sWHI, Hr-Hp, and Wr-Wp were plotted. In the second group, data were stratified by age and sex. Additional stratification by the 5^th^ and 95^th^ sWHI percentiles was conducted to show the sWHI performance in screening abnormal body fat percentage (BFP). Later, data were stratified by the 5^th^ and 95^th^ percentiles of Wt, Wt/Ht, Wt/Ht^2^, and Wt*Ht to evaluate their performance in screening abnormal BFP.

### Descriptive statistics

Descriptive statistics, including percentiles, mean, standard deviation, maximum, and minimum were computed using descriptive functions or self-programming tools. The prctile.m function was used to calculate the 5^th^, 10^th^, 25^th^, 50^th^, 75^th^, 90^th^, and 95^th^ percentiles. The mean.m, median.m, and std.m functions were used to calculate the mean, median, and standard deviation. These statistics explained the key characteristics of the participants and their experimental performance.

### Correlation analysis

The corr.m function was used to calculate the linear or rank correlation between variables. When both variables followed a normal distribution, the Pearson correlation was applied. When either variable did not follow a normal distribution, the Spearman correlation was applied. The jbtest.m function (for Jarque-Bera test) was used to assess normality, with two-tailed p-values ≤0.005 indicating significance [C48]. Mukaka’s guideline was used to interpret correlation coefficients as follows: >0.9 (very high), 0.7 to 0.9 (high), 0.5 to 0.7 (moderate), 0.3 to 0.5 (low) and <0.3 (negligible). Correlation coefficients >0.97 were categorized as extremely high.

### Parametric and non-parametric tests

Continuous variables between two groups were compared using the student’s t-test or Mann-Whitney U test, if approximate, to determine statistical differences [C28].

### Hyperbolic Wt-Ht model

The hyperbolic Wt-Ht model was established through linear regression without introducing the error random variable as follows [C29]:

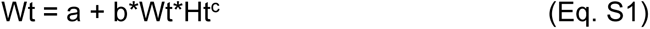

and its transformation:

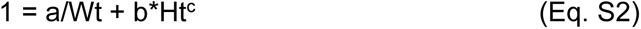

Here, coefficients a and b are sex-, age-, and geography-specific constants. The recommended range for exponent c is -2 to 2, and Wt≠0.

### Wt and Ht prediction models

Based on Eq. 1, we derived a model for predicting Wt based on Ht as follows:

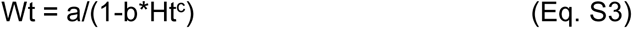

and a model for predicting Ht based on Wt as follows:

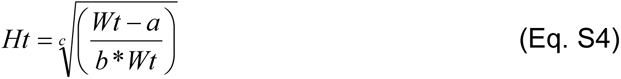

### Regression analysis

We established the hyperbolic Wt-Ht model using linear regression. Furthermore, we established a quadratic polynomial regression model for sex-, age-, and geography- specific coefficients a and b using quadratic regression. The polyfit.m function was used to establish the linear regression model, whereas the fit.m function was used to establish the quadratic polynomial regression model to establish correlation between the independent and dependent variables. For quadratic polynomial regression models, the predint.m function was used to calculate the 95% confidence intervals.

Default values were used for all functions unless otherwise specified.

### Data and code availability

All coding, data processing, and figure plotting were conducted using MATLAB (MathWorks, R2023b, U.S.). We uploaded all source codes and part of the raw data to a GitHub repository (https://github.com/Taukim-Xu/Research-on-Chinese-Hand-Grip-Strength-Biobank/tree/main/Adolescents) to facilitate an in-depth analysis. The source code (except Extended Data Figs. S1 and S2) has been enhanced with detailed comments for readability and is organized by figure sequence numbers.

These source codes and raw data are available for download, use, and modification for non-commercial purposes. The remaining raw data can be obtained by readers from the corresponding data sources.

## Data Availability

Data and code availability
All coding, data processing, and figure plotting were conducted using MATLAB (MathWorks, R2023b, U.S.). We uploaded all source codes and part of the raw data to a GitHub repository (https://github.com/Taukim-Xu/Research-on-Chinese-Hand-Grip-Strength-Biobank/tree/main/Adolescents) to facilitate an in-depth analysis. The source code (except Extended Data Figs. S1 and S2) has been enhanced with detailed comments for readability and is organized by figure sequence numbers. These source codes and raw data are available for download, use, and modification for non-commercial purposes. The remaining raw data can be obtained by readers from the corresponding data sources.

## End notes

## Acknowledgements

We extend our deepest gratitude to all participants for their involvement in this study. We thank the data collection and collation team, including Junfeng Yang, Shanhong Zhou, Jie Peng, Deqiang Lu, Yuhang Guo, Jian Luo, Tao Hu, Fengjia Kang, Jianyin Yan, Junxiong Xu, Zhixian Chen, Jingqi Yong, Mingsen Guo, Xiaoran Wang, Junhao Jiang, Dingfang Wang, and Li Tang. This research was supported by the Research on Science and Technology of Chongqing Municipal Education Commission (No. KJQN202200643), and the General projects of Chongqing Natural Science Foundation of China (No.cstc2020jcyj-msxmX0886). We would like to thank MogoEdit (https://www.mogoedit.com) for its English editing during the preparation of this manuscript.

## Authors contributions

TX directed and contributed to all aspects of this manuscript. WL contributed to data analysis and interpretation. XL, NZ, and QW contributed to the study design. TX wrote the first draft, and WL and XL reviewed and contributed to its modification. TX, XL, NZ, and QW contributed to the data collection. All authors read and approved the final manuscript, and provided feedback.

## Competing interests

The authors declare that they have no competing interests.

## Additional Information

Correspondence and requests for materials should be addressed to Taukim Xu through.

## Abbreviations

Wt: Weight
Ht: Height
mHGS: maximum Hand Grip Strength
BMI: Body Mass Index
IBW: Ideal Body Weight
sWHI: standardized Weight-Height Index
BFP: Body Fat Percentage
BCCPD: Body Composition of Chinese People Data Set
KSPF: Korea Sports Promotion Foundation
NHANES: National Health and Nutrition Examination Survey
JDHLE: Japanese Database for Human Life Engineering
WHO: World Health Organization
Hr-Hp: difference between predicted Height and real Height
Wr-Wp: difference between predicted Weight and real Weight
Hr: real Height
Hp: predicted Height
Wr: real Weight
Wp: predicted Weight
std.: standard deviation
CI: Confidence Interval

## Extended Data

**Extended Data Fig. S1:**
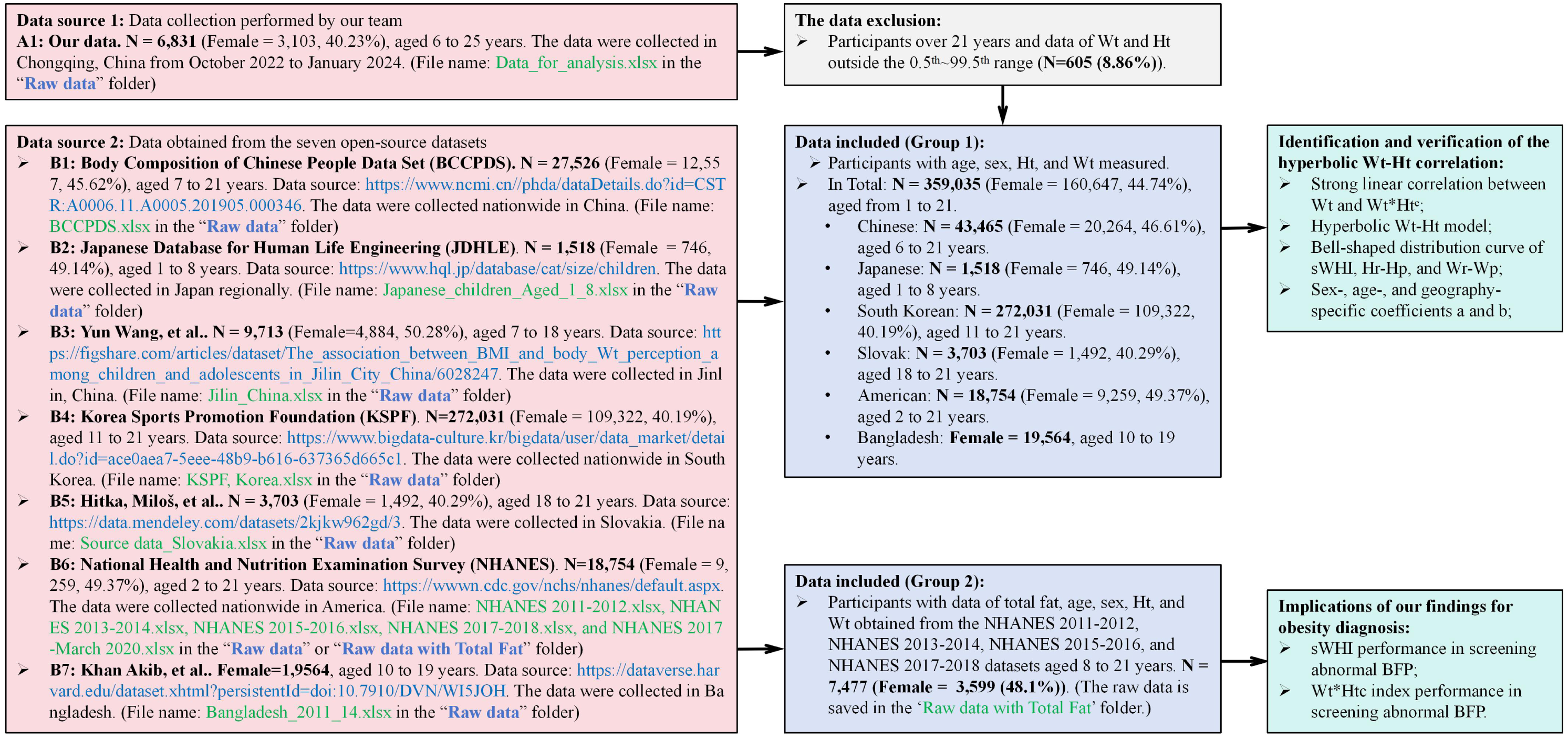
Summary of data collection, data inclusion and exclusion, and data flow. BMI = Body Mass Index; sWHI = standardized weight-height index; BFP = body fat percentage; Wt = weight; Ht = height; Hr = real height; Hp = predicted height; Wr = real weight; Wp = predicted weight; BCCPDS = Body Composition of Chinese People Data Set; JDHLE = Japanese Database for Human Life Engineering; KSPF = Korea Sports Promotion Foundation; NHANES = National Health and Nutrition Examination Survey

**Extended Data Fig. S2:**
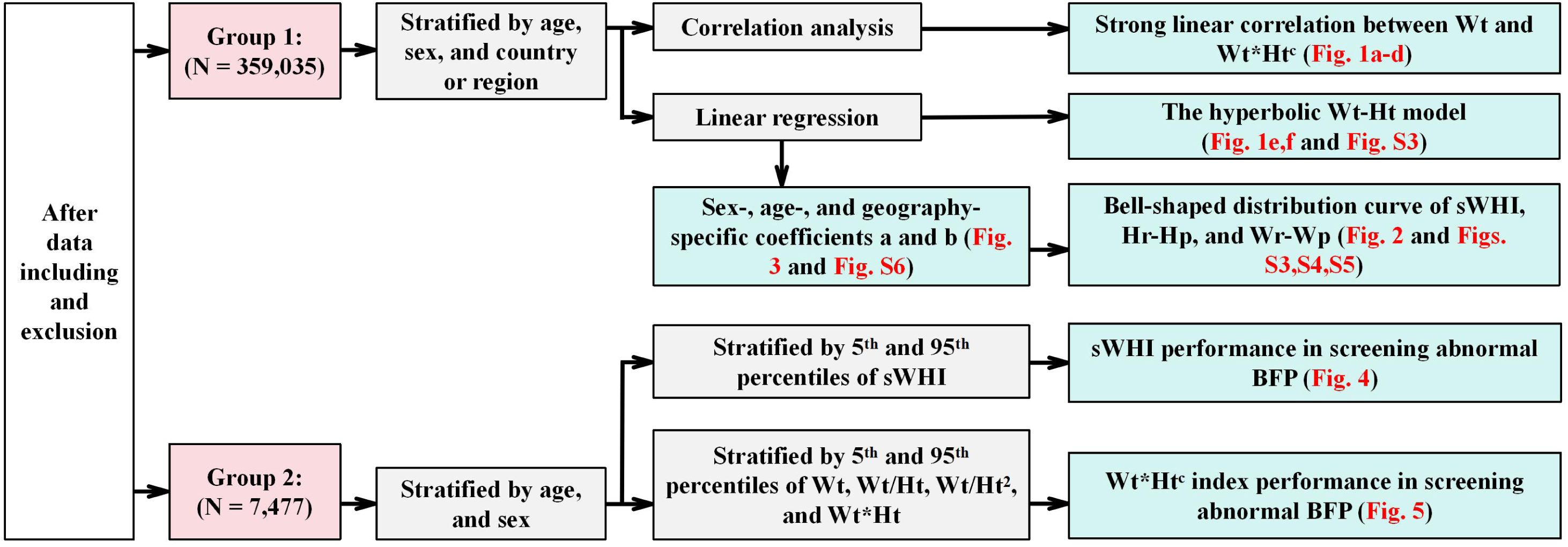
Summary of data flow. sWHI = standardized weight-height index; BFP = body fat percentage; Wt = weight; Hr = real height; Hp = predicted height; Wr = real weight; Wp = predicted weight

**Extended Data Fig. S3:**
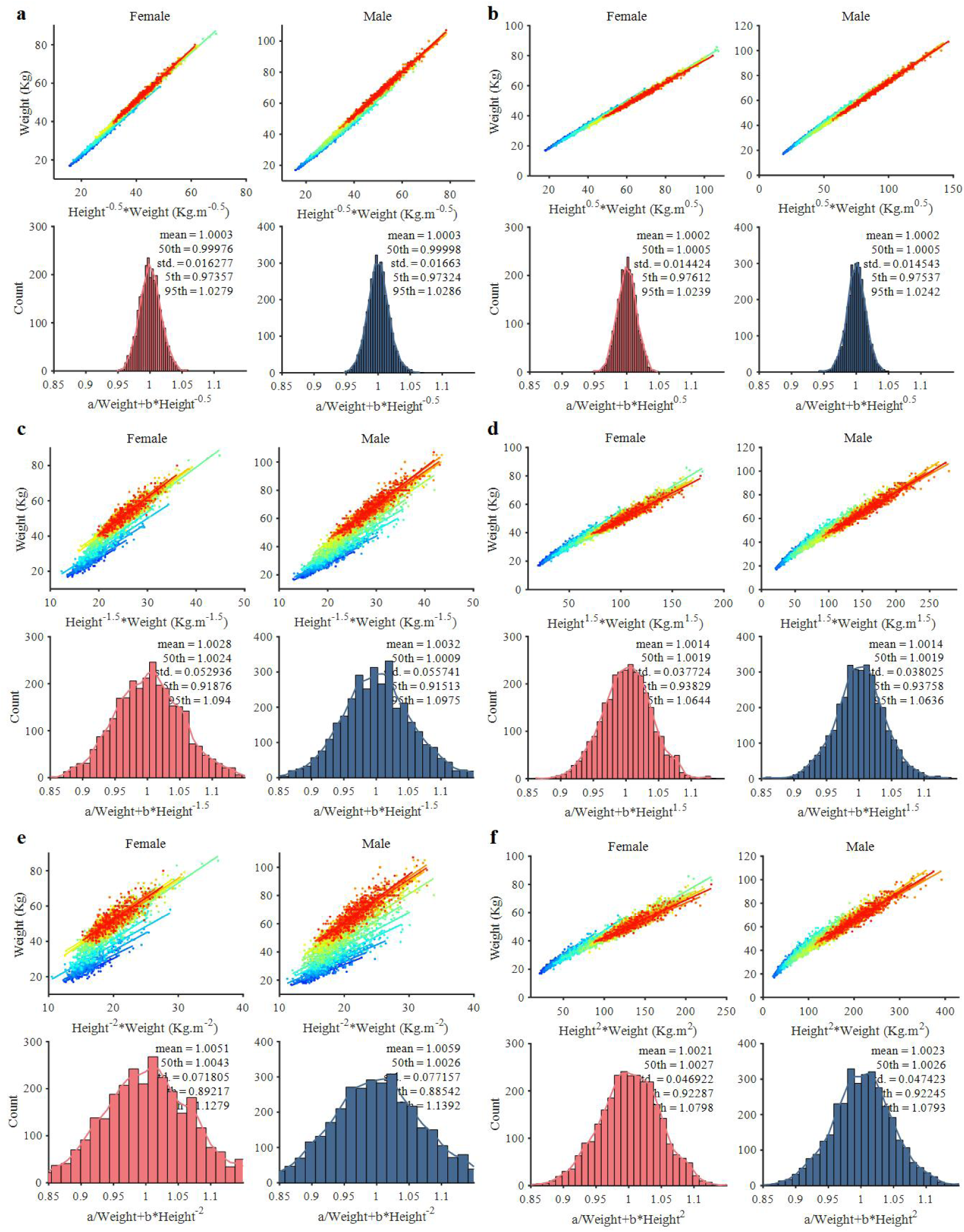
Linear regression of the hyperbolic Wt-Ht model and sWHI distributions when the exponent c ranges from -2 to 2. **a-f**, Hyperbolic Wt-Ht model and sWHI distributions when the exponent c=-0.5 (**a**), 0.5 (**b**), -1.5 (**c**), 1.5 (**d**), -2 (**e**), and 2 (**f**). Mean, standard deviation, and 5^th^, 50^th^, and 95^th^ percentiles are shown for all sWHI distributions. Wt = weight; Ht = height; sWHI = standardized weight-height index; std. = standard deviation

**Extended Data Fig. S4:**
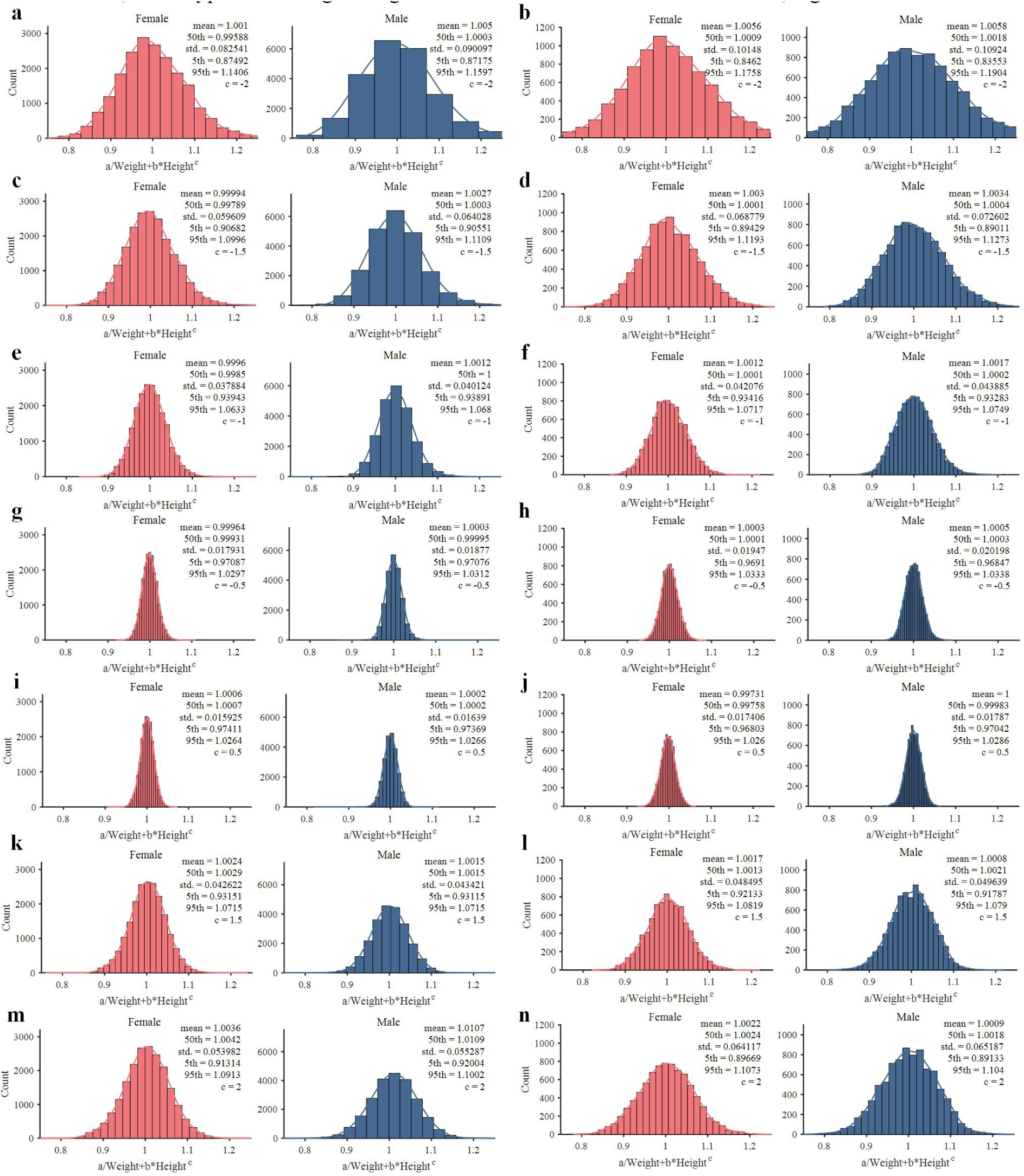
sWHI distributions when the exponent c ranges from -2 to 2 for all Chinese and American datasets. **a-n**, sWHI distributions for all Chinese (**a,c,e,g,i,k,m**) and American (**b,d,f,h,j,l,n**) datasets, when the exponent c=-2 (**a,b**), -1.5 (**c,d**), -1 (**e,f**), -0.5 (**g,h**), 0.5 (**i,j**), 1.5 (**k,l**), and 2 (**m,n**), respectively. Mean, standard deviation, and 5^th^, 50^th^, and 95^th^ percentiles are shown for all sWHI distributions. sWHI = standardized weight-height index; std. = standard deviation

**Extended Data Fig. S5:**
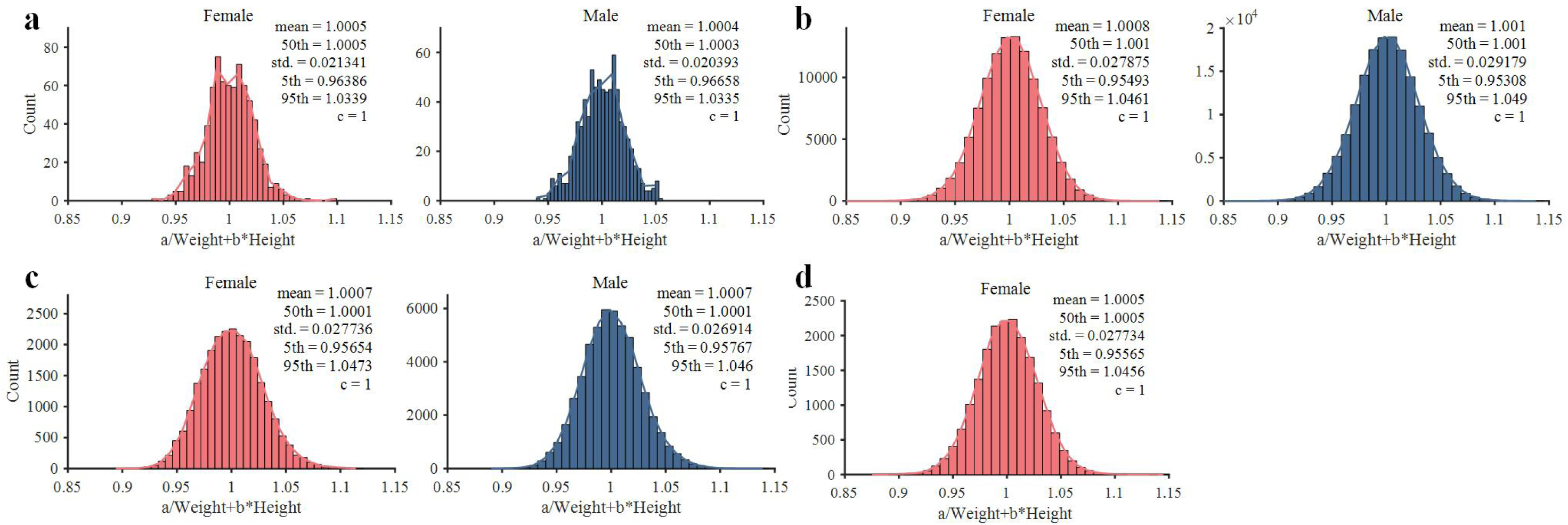
sWHI distributions when the exponent c=1 across other datasets. **a-d**, sWHI distributions for B2: JDHLE. (**a**), B4: KSPF (**b**), B5: Hitka, Miloš, et al. (**c**), and B7: Khan Akib, et al. (**d**). Mean, standard deviation, and 5^th^, 50^th^, and 95^th^ percentiles are shown for all sWHI distributions. sWHI = standardized weight-height index; std. = standard deviation; JDHLE = Japanese Database for Human Life Engineering; KSPF = Korea Sports Promotion Foundation

**Extended Data Fig. S6:**
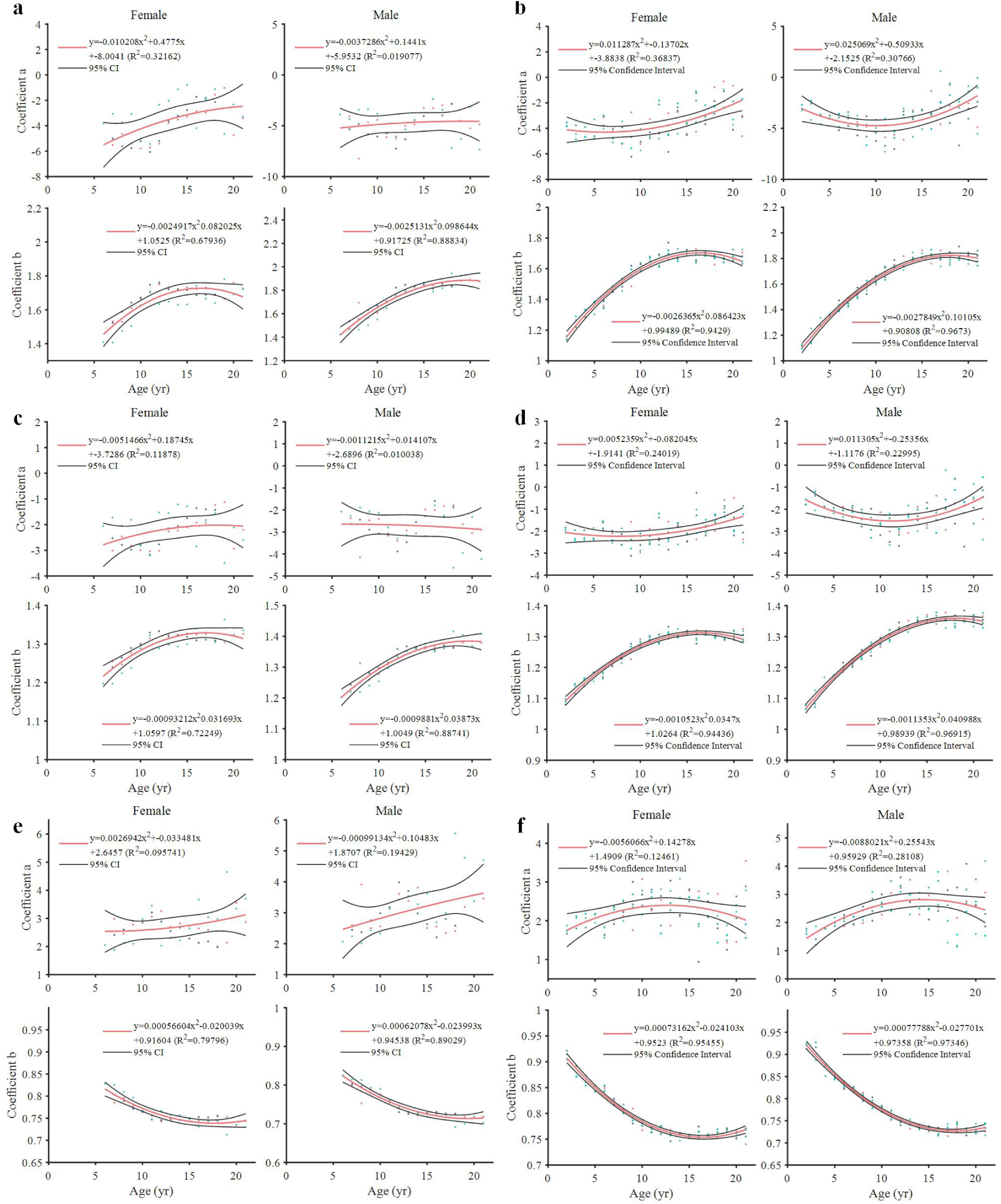
Quadratic polynomial model for coefficients a and b when the exponent c=-1, -0.5, and 0.5 for all Chinese and American datasets. **a-f**, Quadratic polynomial model for coefficients a and b for all Chinese (**a,c,e**) and American (**b,d,f**) datasets when the exponent c=-1 (**a,b**), -0.5 (**c,d**), and 0.5 (**e,f**). R^2^ = determination coefficient in regression analysis; CI = confidence interval

**Extended Data Fig. S7:**
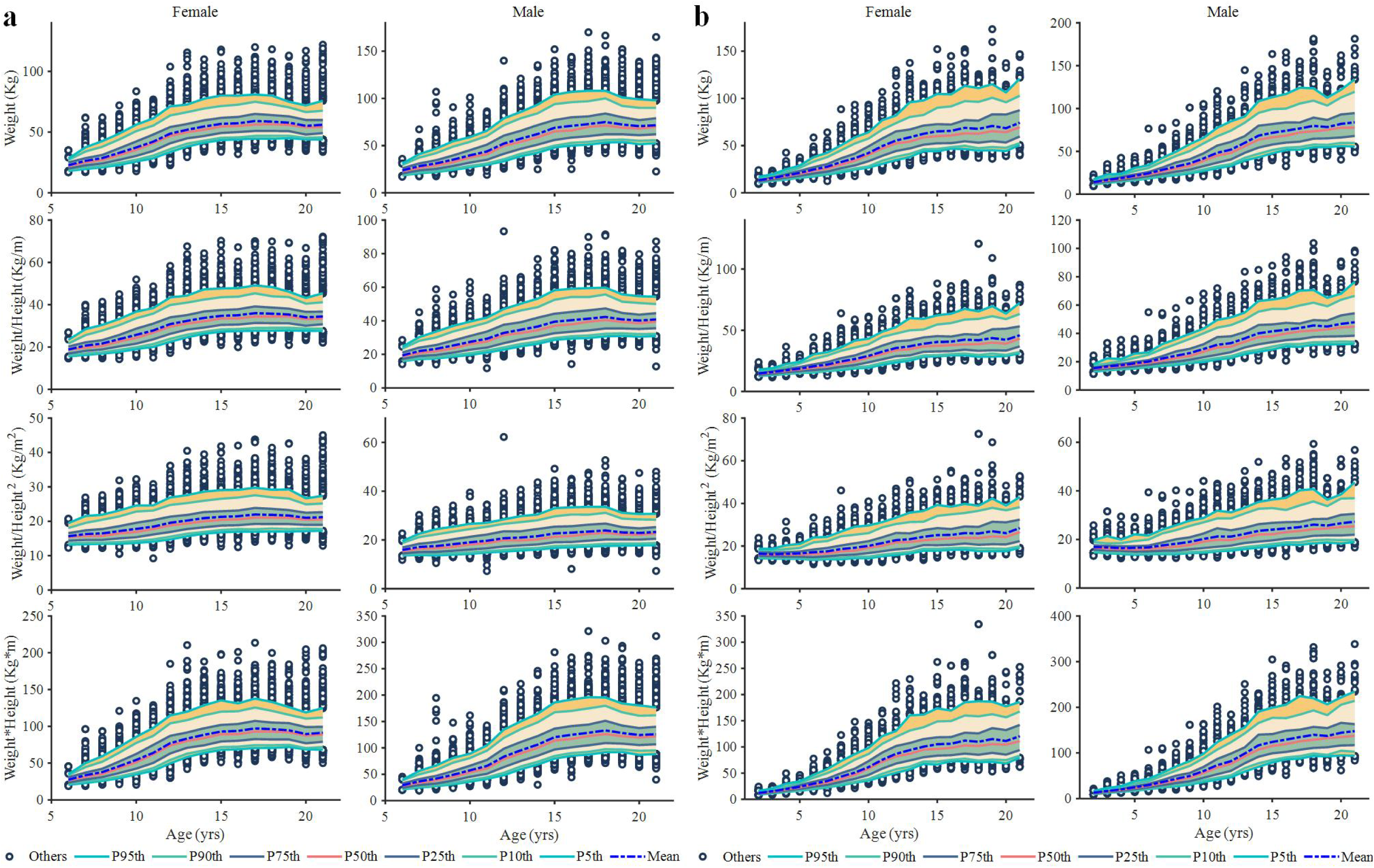
Wt*Ht^c^ indexes percentiles when the exponent c=0, -1, -2, and 1 for all Chinese and American datasets. **a,b,** Percentiles of Wt, Wt/Ht, Wt/Ht^2^, and Wt*Ht for all Chinese (**a**) and American (**b**) datasets. Wt = weight; Ht = height

**Extended Data Fig. S8:**
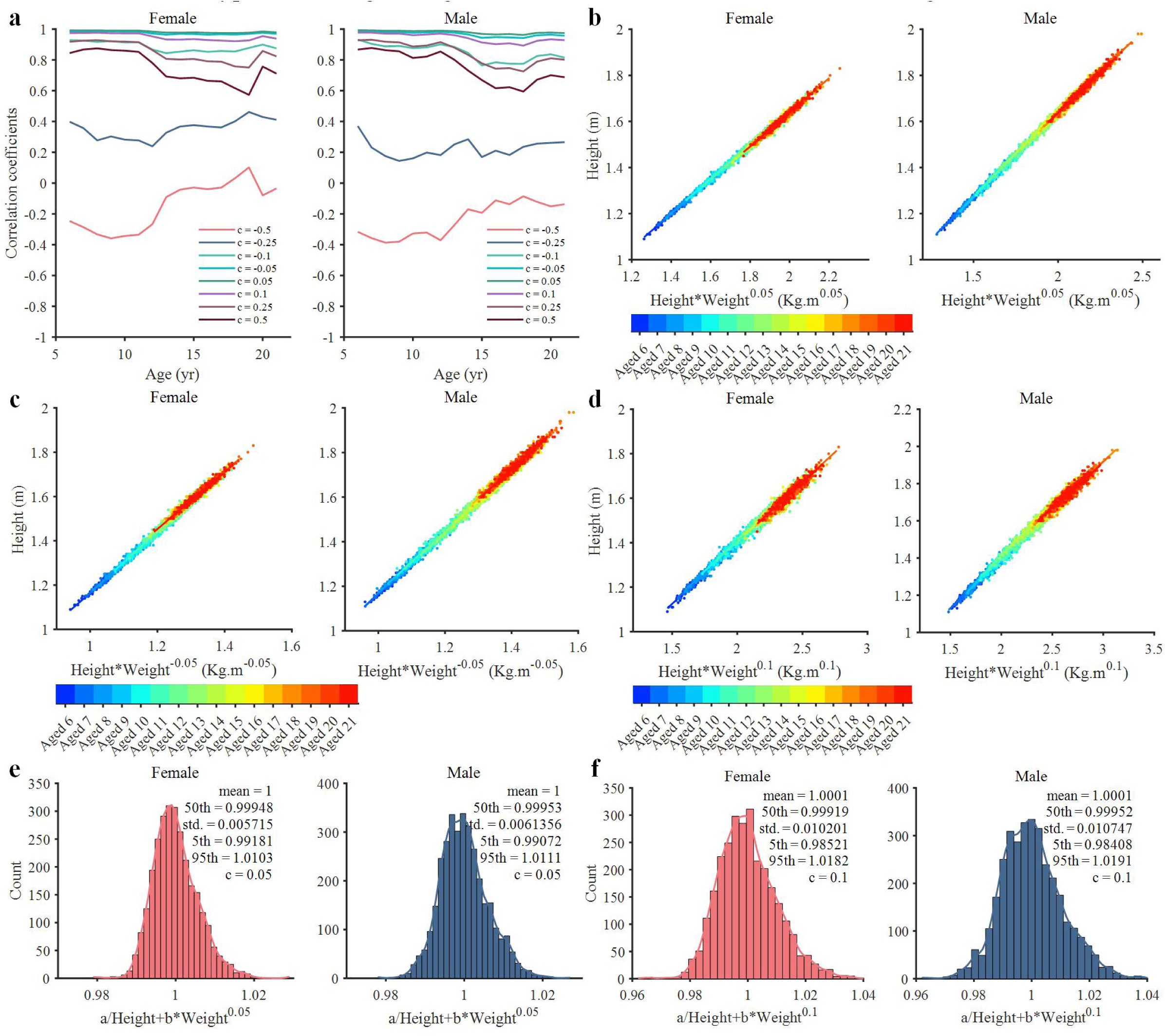
Hyperbolic Wt-Ht model formulated as Ht≈Ht*Wt^c^ for all Chinese datasets. **a**, Correlation coefficients between Ht and Ht*Wt^c^ when the exponent c ranges from -0.5 to 0.5. **b-d**, Linear regression of Ht=a+b*Ht*Wt^c^ when the exponent c=0.05 (**b**), -0.05 (**c**), and c=0.1 (**d**). **e,f**, Distributions of a/Ht+b*Wt^c^ when the exponent c=0.05 (**e**) and 0.1 (**f**). Mean, standard deviation, and 5^th^, 50^th^, and 95^th^ percentiles are shown for all sWHI distributions. Wt = weight; Ht = height; std. = standard deviation

**Extended Data Fig. S9:**
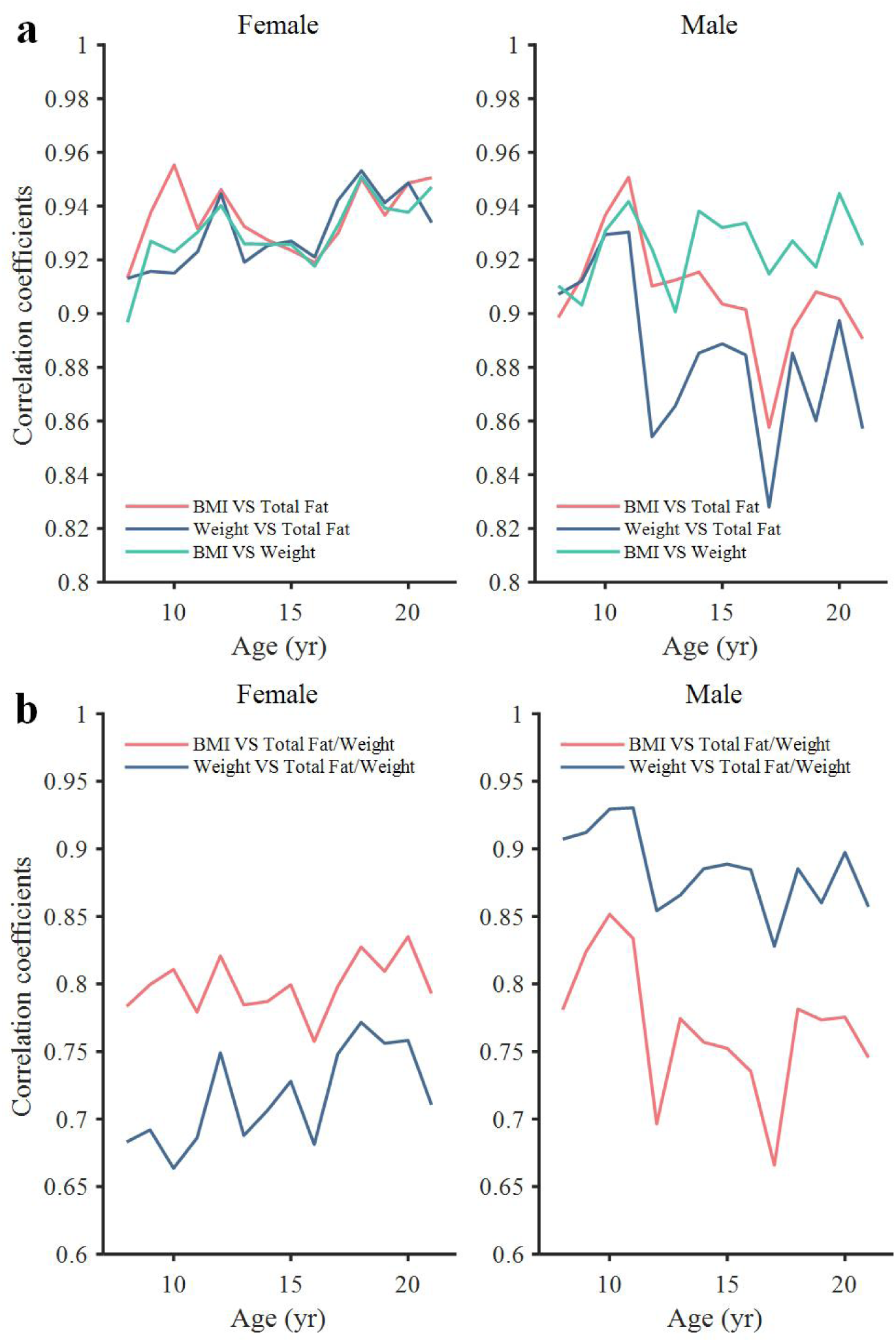
Correlation coefficients between Wt, BMI, body total fat, and BFP. **a,b,** Correlation coefficients between BMI and total fat, Wt and total fat, and BMI and Wt (**a**), and between BMI and BFP, and Wt and BFP (**b**) based on date from the NHANES. BMI = body mass index; BFP = total fat/weight; Wt = weight; NHANES = National Health and Nutrition Examination Survey

## Notes

### Competing Interest Statement

The authors have declared no competing interest.

### Author Declarations

This study was approved by the Institutional Review Board of the People's Hospital of Chongqing Banan District (approval number BNLL-KY-2023-004).

